# Burden of five healthcare associated infections in Australia

**DOI:** 10.1101/2021.10.05.21264587

**Authors:** M. J. Lydeamore, B.G. Mitchell, T. Bucknall, A.C. Cheng, P.L. Russo, A. J. Stewardson

## Abstract

Healthcare associated infections (HAIs) are associated with increased morbidity and mortality, but there are few data that quantify the burden of infection nationally in Australia. We used data from an Australian national point prevalence survey to estimate the burden of HAIs amongst adults in Australian public hospitals. The incidence based appropach, introduced by the ECDC Burden of Comunicable Diseases in Europe was used. We estimate that 170,574 HAIs occur in adults admitted to public hospitals in Australia annually, resulting in 7583 deaths. Hospital acquired pneumonia is the most frequent HAI, followed by surgical site infections, and urinary tract infections. We find that blood stream infections contribute a small percentage of HAIs, but contribute the highest number of deaths (3512), more than twice that of the second largest, while pneumonia has the highert impact on years lived with disability. This study is the first time the national burden of HAIs has been estimated for Australia from point prevalence data. The estimated burden is high, and highlights the need for continued investment in HAI prevention.

## 1. Introduction

Healthcare associated infections (HAIs) are associated with increased morbidity and mortality, and excess healthcare costs (1,2). An accurate quantification of HAI burden is required to prioritise and evaluate infection prevention interventions. The burden of HAIs overseas is known to be high (3–8), but previous estimates in an Australian setting have relied on a range of opportunistic reports of incidence (9).

The ECDC introduced a methodology to estimate the total number of HAI cases from a point prevalence survey (4). Combined with disease outcome trees, the number of disability adjusted life years (DALYs) and deaths can also be estimated (10).

We previously conducted a point prevalence survey (PPS) of HAIs across 19 public hospitals in Australia (11). We used data from this PPS, combined with admitted patient care data from the Australian Institute of Health and Welfare (12). The aim was to estimate the population level burden in Australia of five HAIs: healthcare-associated *Clostridioides difficile* infection (CDI), healthcare-associated bloodstream infection (BSI), urinary tract infection (UTI), healthcare acquired pneumonia (HAP) and surgical site infection (SSI). These estimates are also compared with previous literature-based estimates of the burden of HAIs in Australia, and with similar PPS studies conducted in Europe.

## 2. Methods

### 2.1 Study population and study design

The Australian PPS data used in this study was collected in 2018 in a sample of adult patients in 19 public, large acute care hospitals. The surveillance methodology was based on the European Centre for Disease Prevention and Control (ECDC) PPS protocol (13). The types of HAIs that were selected for this study were as described in Cassini et. al (4). HAIs were defined as per the ECDC protocol (13), with data collected by two research assistants, and entered into a secure online web-based survey tool.

A total of 2767 patients were sampled. Results from this PPS have previously been reported in detail (11,14). The median age of patients was 67 (IQR 49-79, range 18-104). Of these, 52.9% (1465) were male, 46.6% (1289) female and 0.5% (13) unknown/other. A majority (85.7%) of patients were from major city hospitals, with the remaining 14.3% from regional services.

### 2.2 Outcome measures

As well as the number of cases, we estimate deaths and DALYs for each condition. DALYs are a composite measure of years lived with disability (YLDs) and years life lost (YLLs), accounting for incidence, severity, and mortality of disease simultaneously. They also provide a way to compare the impact of disease across conditions, as opposed to simply ranking by incidence of prevalence.

### 2.3 Estimation methodology

The same approach as used for the estimation of the burden of HAIs in Germany was applied to this PPS (3), except for the choices of age strata. As the Australian PPS was only collected in adults, and involved a smaller sample size, strata were chosen to be 18-24, 25-34, 35-44, …, >75. As the probability of death following an SSI is dependent on age (and thus on strata), the BHAI R package was modified to be compatible with these strata (Personal communication, B. Zacher). The disease outcome trees, transition probabilities and disability weights were otherwise the same as though used by Cassini et al. (4). For full details of the outcome trees, see the supplement of Cassini et al. or the ECDC BCoDE toolkit (10).

The process of estimation can be summarised into three steps. The first step is to use the PPS data to estimate the hospital prevalence, which is estimated as

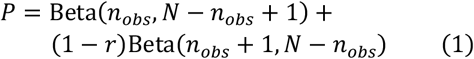

where *n*_*obs*_ is the number of patients observed with a HAI and *N* is the total number of patients in the PPS. Next, this estimate is converted to hospital incidence,

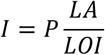

where *P* is the hospital prevalence from Eq. (1), *LA* is the mean length of stay and *LOI* is the mean length of infection. For this study, the mean length of stay, *LA*, was set to 5.3 days, from the AIHW 2018 statistics on all public hospitals (excluding same-day separations). Following the methodology of Zacher et. al, the mean length of infection was estimated using the censored length of infection from the survey and the Grenander estimate.

The final step in the estimation is the hospital incidence, which is calculated as

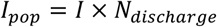

The survey used in this study was in acute public hospitals for patients over 18 years of age, which accounts for approximately 60% of separations in public hospitals for patients over 18 years of age, giving *N*_*discharge*_ = 3,713,513.

To enable comparison between the European and German burden estimates, both datasets were re-aggregated to match the wider stratification used for estimation in the Australian setting. It is noted that the data for these surveys is aggregated into five-year age bands, and so the lowest age category for these studies is 15-24 (as opposed to 18-24). However, the burden in those aged between 15 and 18 is relatively low, so is expected to have little impact on the results.

As the Australian PPS used the ‘light’ survey design as specified by the ECDC, McCabe scores are not recorded. We applied the McCabe score distribution of the ECDC PPS to Australia, assuming that the McCabe score distribution in Australia would be similar to that observed in the EU. It is noted that there is little evidence of the applicability or lack thereof of these estimates to the Australian population.

### 2.4 Ethical Statement

For the initial PPS study, ethical approval was granted by the Alfred Health Human Research Ethics Committee (HREC/17/Alfred/203) through the Australian National Mutual Assessment process for all states and territories except for Tasmania, for which a separate approval was obtained from the Tasmanian Health and Medical Human Research Committee (H0016978). This study was based on health information collected and published within the Australian PPS or published within the ECDC PPS and did not require informed consent from participants. Data were provided in aggregated form by age and sex strata, with no personal identifiers.

## 3. Results

### 3.1 Burden of healthcare-associated infections in Australia

The estimates for the total number of cases of HAIs is contained in Table 1. It is estimated that there are approximately 7500 deaths each year from HAIs in Australia, with the majority being caused by blood stream infections (BSI). More than 122,000 DALYs are contributed by HAIs, with the two largest contributors being BSI and healthcare-acquired pneumonia (HAP).

**Table 1:** Annual burden of five healthcare associated infections (HAIs), estimated from Australian point prevalence survey data from 2018. Numbers inside brackets indicate 95% uncertainty intervals (UI). SSI, surgical site infections; UTI, urinary tract infections; CDI, *Clostridioides difficile* infection; HAP, healthcare acquired pneumonia; BSI, bloodstream infection; DALYs, disability adjusted life years; YLL, years of life lost; YLD, years lived with disability.

|  | **Number of HAIs**<br>**(95% UI)** | **Deaths**<br>**(95% UI)** | **DALYs**<br>**(95% UI)** | **YLL**<br>**(95% UI)** | **YLD**<br>**(95% UI)** |
| --- | --- | --- | --- | --- | --- |
| **SSI** | 44,238<br>(31,176 - 63,797) | 876<br>(617 - 1,263) | 13,197<br>(9,298 - 19,001) | 12,982<br>(9,149 - 18,722) | 214<br>(145 - 317) |
| **UTI** | 42,408<br>(25,200 - 68,735) | 729<br>(259 - 1,772) | 16,087<br>(5,939 - 37,218) | 10,983<br>(3,899 - 26,704) | 4,879<br>(1,745 - 11,659) |
| **CDI** | 5,125<br>(2,360 - 10,740) | 262<br>(13 - 836) | 2,757<br>(241 - 8,655) | 2,635<br>(128 - 8,403) | 127<br>(21 - 384) |
| **HAP** | 51,499<br>(31,343 - 82,877) | 1,904<br>(462 - 4,430) | 39,276<br>(17,608 - 77,915) | 23,245<br>(5,644 - 54,078) | 15,684<br>(8,038 - 28,817) |
| **BSI** | 23,979<br>(15,658 - 36,245) | 3,512<br>(1,874 - 6,075) | 46,773<br>(26,205 - 79,104) | 39,665<br>(21,159 - 68,610) | 6,964<br>(3,660 - 12,446) |
| **All** | 170,574<br>(135,779 - 213,898) | 7,583<br>(4,941 - 11,135) | 122,376<br>(85,136 - 172,784) | 93,322<br>(61,443 - 135,722) | 28,669<br>(18,571 - 43,924) |

As is expected, the incidence of HAIs appears inherently age-based (Figure 1), with those aged greater than 75 having 21 times more cases than those aged 18 to 24. This is consistent across incidence, DALYs (Figure 2) and attributable deaths (Figure 3).

**Figure 1:**
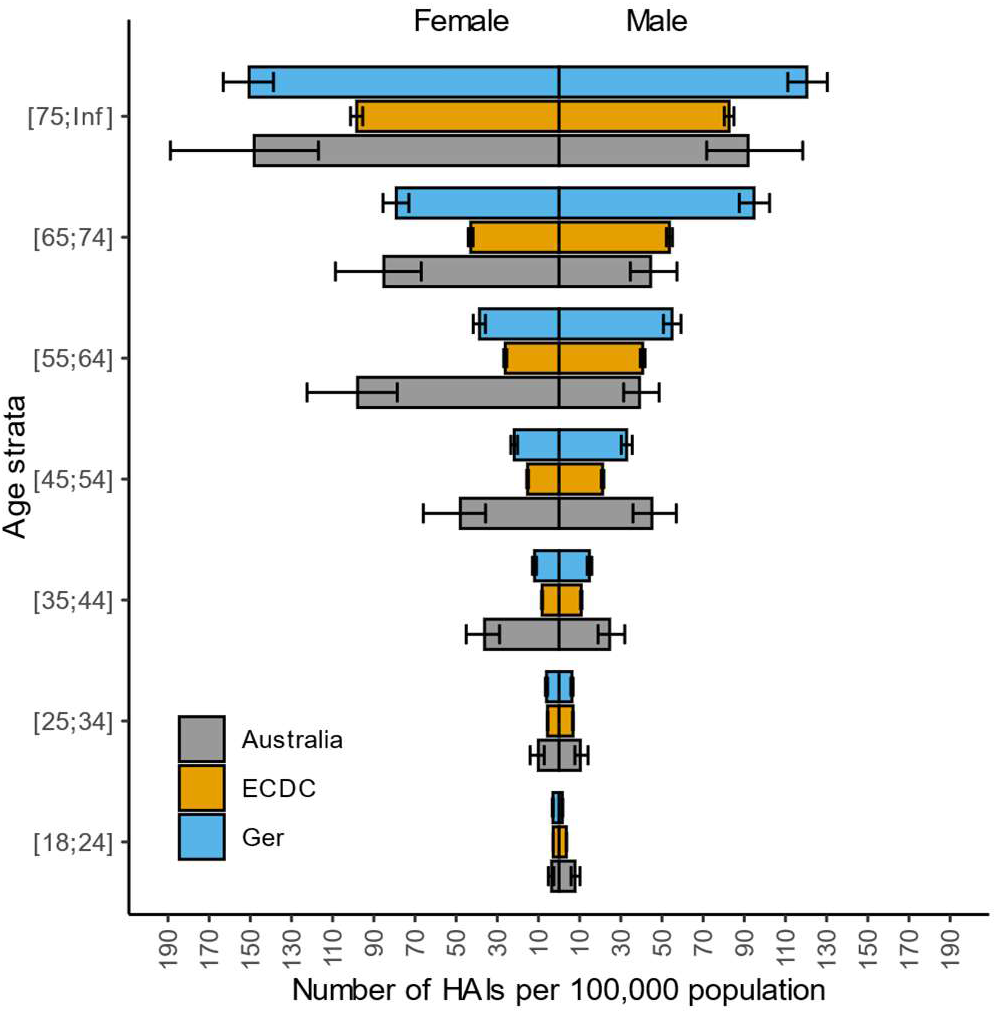
Number of cases of healthcare associated infections (HAIs) per 100,000 population, stratified by age and sex, in Australia, the Europe and Germany.

**Figure 2:**
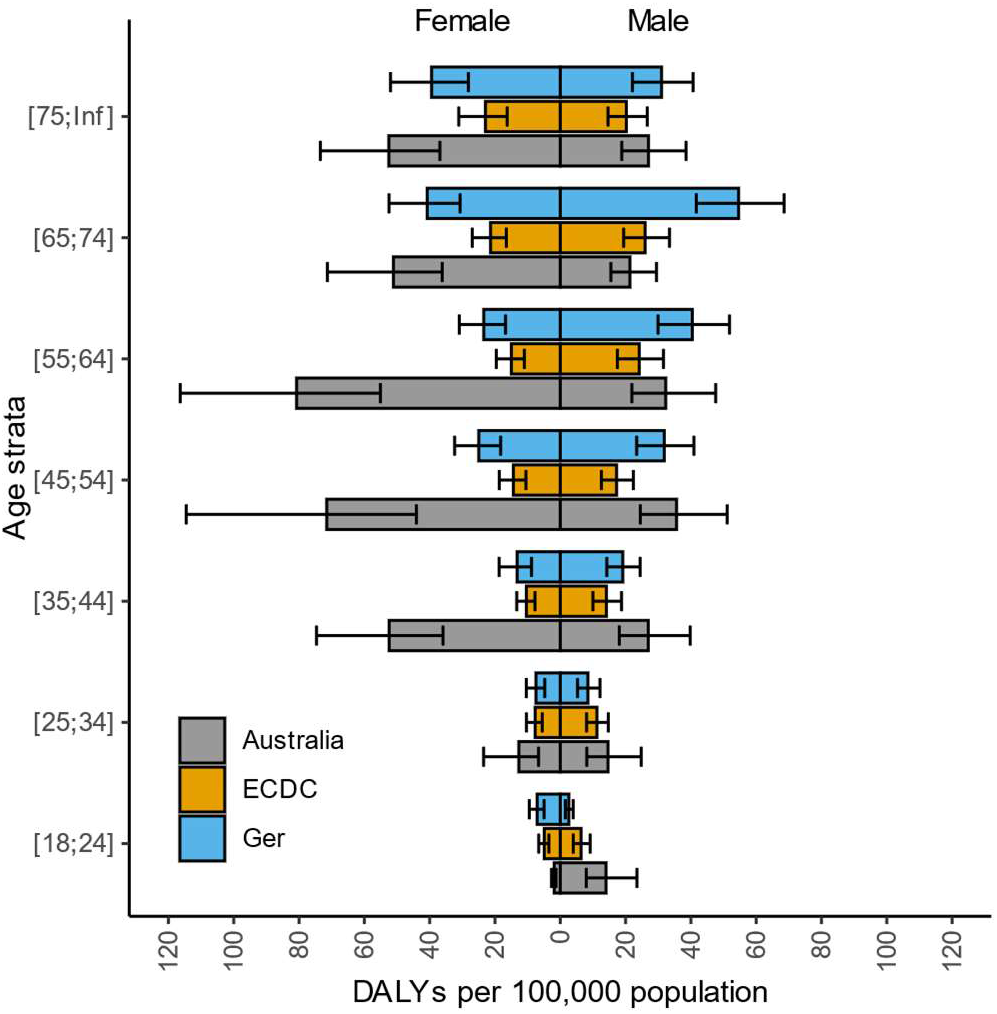
Number of disability adjusted life years (DALYs) from healthcare associated infections per 100,000 population, stratified by age and sex, in Australia, the Europe and Germany

**Figure 3:**
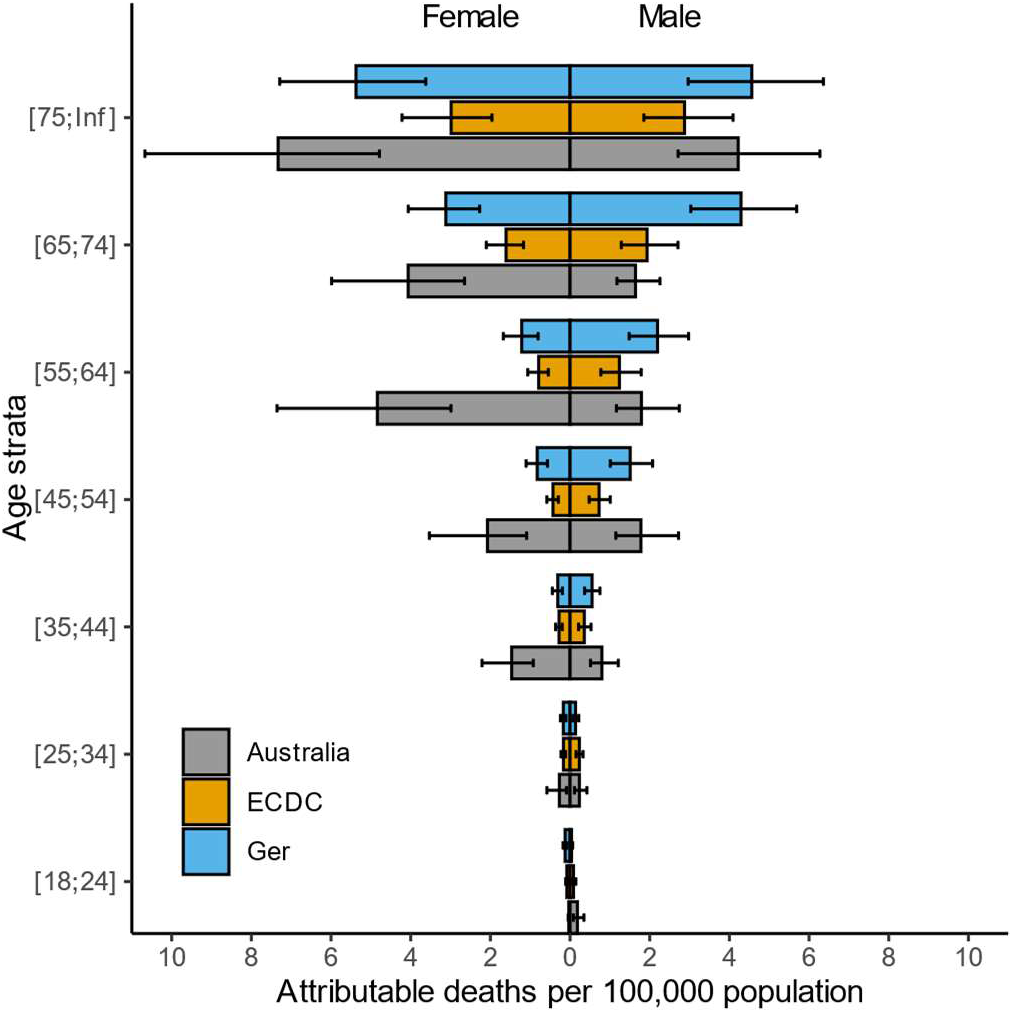
Number of attributable deaths from healthcare associated infections per 100,000 population, stratified by age and sex, in Australia, Europe and Germany.

### 3.2 Comparison with global estimates

Burden per 100,000 population for Australia, the EU and Germany are given in Table 2 and shown in Figure 1. Overall, the estimated burden in Australia is higher compared to both the EU and Germany across all age categories, and particularly so in females aged between 55 and 64. When considered by HAI type (Figure 4) shows that the rate of healthcare acquired pneumonia and bloodstream infection are significantly higher than that of the other two settings. Similar to international observations, the rate of UTI is higher in females, although a similar trend is observed in Australian HAP and BSI numbers compared to international estimates.

**Table 2:** Annual burden per 100,000 population of five types of healthcare-associated infections from the Australian PPS, and the ECDC PPS Sample. SSI, surgical site infections; UTI, urinary tract infections; CDI, *Clostridioides difficile* infection; HAP, healthcare acquired pneumonia; BSI, bloodstream infection; DALYs, disability adjusted life years; YLL, years of life lost; YLD, years lived with disability.

|  | Sample | SSI | UTI | CDI | HAP | BSI | All |
| --- | --- | --- | --- | --- | --- | --- | --- |
| HAIs | Australia | 179.8<br>(126.7 - 259.3) | 172.4<br>(102.4 - 279.4) | 20.8<br>(9.6 - 43.7) | 209.3<br>(127.4 - 336.9) | 97.5<br>(63.7 - 147.3) | 693.4<br>(551.9 - 869.5) |
|  | ECDC | 19.9<br>(17.7 - 22.2) | 128.7<br>(122.4 - 135.1) | 155.9<br>(148.6 - 163.5) | 99.4<br>(94.3 - 104.6) | 14.4<br>(12.7 - 16.4) | 418.4<br>(407.3 - 430) |
|  | German | 162.8<br>(137.6 - 191) | 146<br>(127.1 - 167.8) | 52.6<br>(41.9 - 65.8) | 228.8<br>(199.7 - 262.6) | 44.6<br>(35.6 - 55.3) | 636.3<br>(589 - 686.7) |
| Deaths | Australia | 3.6<br>(2.5 - 5.1) | 3<br>(1.1 - 7.2) | 1.1<br>(0.1 - 3.4) | 7.7<br>(1.9 - 18) | 14.3<br>(7.6 - 24.7) | 30.8<br>(20.1 - 45.3) |
|  | ECDC | 3<br>(1.9 - 4.1) | 4.8<br>(1.2 - 9.2) | 2.7<br>(1.1 - 5.4) | 2.3<br>(2.2 - 2.4) | 0.8<br>(0 - 1.6) | 13.8<br>(9.3 - 19.1) |
|  | German | 6.1<br>(1.6 - 12) | 3.7<br>(3.2 - 4.2) | 7.8<br>(4.8 - 11.6) | 4<br>(1.6 - 8) | 2.4<br>(0.1 - 5.1) | 24.5<br>(16.9 - 32.7) |
| DALYs | Australia | 53.6<br>(37.8 - 77.2) | 65.4<br>(24.1 - 151.3) | 11.2<br>(1 - 35.2) | 159.7<br>(71.6 - 316.7) | 190.1<br>(106.5 - 321.6) | 497.5<br>(346.1 - 702.4) |
|  | ECDC | 43.1<br>(29.1 - 58.1) | 84.4<br>(43.7 - 131.1) | 48.4<br>(20.6 - 88.2) | 30.3<br>(28.8 - 31.9) | 8.3<br>(0.7 - 16.2) | 216.5<br>(158.8 - 280.9) |
|  | German | 98.6<br>(51 - 159.3) | 44.6<br>(38.8 - 51.3) | 102.2<br>(66.2 - 148.2) | 70.3<br>(30.1 - 130.8) | 23.8<br>(2.1 - 49) | 344.7<br>(256 - 442.2) |

**Figure 4:**
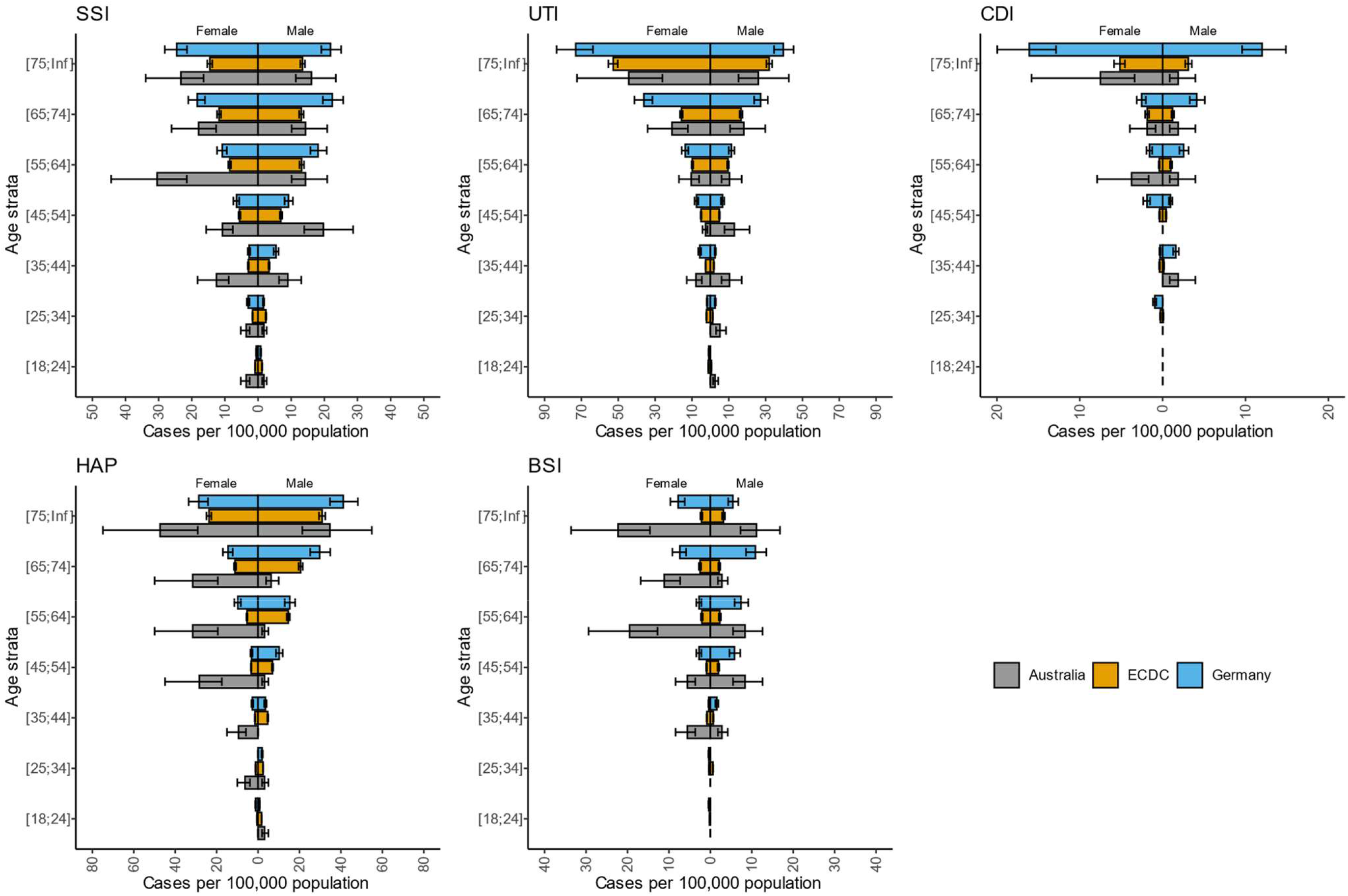
Number of healthcare acquired infections, stratified by age, sex and infection type, normalised by population, in Australia, Europe and Germany. SSI, surgical site infections; UTI, urinary tract infections; CDI, *Clostridioides difficile* infection; HAP, healthcare acquired pneumonia; BSI, bloodstream infection.

In all estimates, the uncertainty for the Australian population is much higher than that for the EU and Germany. This is likely due to the relatively small number of patients in this PPS (2767), as opposed to 273,753 patients in the ECDC survey and 41,539 in the German convenience sample.

### 3.3 Comparison to other conditions

At a rate of 498 DALYs per 100,000 population, HAIs contribute substantially to the overall burden of disease in Australia. For comparison, it is estimated that motor vehicle injuries contribute 180 DALYs, infectious diseases 370 DALYs and respiratory diseases 1380 DALYs per year in 2015 (15). While substantially less in magnitude than cancer and other neoplasms – which contribute 2400 DALYs annually – the health savings on these largely preventable conditions are substantial in Australia.

## 4. Discussion

We have estimated the burden of five HAIs in Australia based on point prevalence data from 2018. By computing the number of DALYs, we have provided a comparison point for other health conditions in Australia. We have also compared these estimates to similar studies in Germany and the EU.

Previous estimates of the burden of HAIs in Australia were based on combinations of studies with highly varied collection protocols (9) or based on a study in a single hospital (16). A previous review of these literature reported that the burden of HAIs was approximately 83,000 per year, almost half of that estimated here (9). However, the study noted incomplete data on pneumonia and bloodstream infections, and if that data were complete, the incidence may be closer to 165,000 per year, similar to our estimate.

In comparison to the EU and Germany, the incidence of HAIs in Australia is significantly larger, although the uncertainty on our estimates is comparatively high. In these two settings, the largest contributors to the HAI burden are UTIs, HAP and SSIs, similar to Australia. Interestingly, HAP contributes the most DALYs in the EU estimates, compared with BSI in both Germany and Australia.

There are potential methodological explanations for the higher incidence estimate in Australia. First, the Australian point prevalence survey was performed in large acute public hospitals, whereas the European surveys were performed in a wider range of facilities. Second, the number of separations used for the EU estimates is an approximation derived from the number of patient-days, whereas we have the actual total number of separations, but have assumed the percentage of these that were in adults and in public, acute hospitals.

The findings in this study are subject to the same limitations of the source PPS study, including selection bias, restriction to public hospitals and the lack of patient-level factors present in the data. Although we have used a very similar methodology to other studies overseas, these limitations mean that direct comparison between studies is challenging.

We are also limited by the number of sample patients. Although 2767 patients is substantially more than a number of single site studies in the same setting, it is orders of magnitude less than the German and EU point prevalence surveys. The impact of this limitation is that the uncertainty in our estimates is relatively high, particularly when stratified by even broad age bands.

We utilised the McCabe score distribution from the European point prevalence survey, as the Australian PPS used the “light” definition and so did not collect its own scores. There is a large number of “avoidable” hospital stays in Australia, even in the acute hospitals that were the focus of the PPS. Therefore, the use of the European McCabe scores would overestimate the severity of underlying disease, and thus underestimate the DALYs, as assumed remaining life expectancies would be lower.

HAIs are a significant public health issue in Australia when compared with other health conditions. Our findings are consistent with HAI estimates in European settings. Of note, is the large contributions that pneumonia and BSI have on overall burden and in particular deaths and DALYs. This study shows the need for continued investment in the prevention of HAIs in Australia, and importantly for robust, national coordinated surveillance of these conditions.

## Data Availability

Data requests should be made by email to the corresponding author (M. J. Lydeamore)

## Acknowledgements

The authors would like to thank Dr B. Zacher for his help in modifying the BHAI R package to function with our data format.

AJS and PLR are supported by an NHMRC Early Career Fellowship (GNT1141398 and APP1156312 respectively). MJL is supported by an NHMRC Project Grant (GNT1156742). BGM is a recipient of a NHMRC Investigator Grant (2021-2026; APP2008392).

